# Mindfulness facets and psychological well-being among meditators: Serenity as a mediating process

**DOI:** 10.1101/2023.09.17.23295683

**Authors:** Rebecca Y. M. Cheung, Iris Yili Wang, Elsa Ngar-Sze Lau

## Abstract

Guided by the theoretical processes of mindfulness and psychological well-being, this study examined serenity as a mediator between mindfulness facets and psychological well-being, as indexed by depressive symptoms and life satisfaction. Participants were 133 mindfulness practitioners who took part in a 3-day transnational meditation event in Hong Kong. Upon informed consent, participants completed a self-report questionnaire. The findings from structural equation modeling showed that serenity mediated the relation between two facets of mindfulness, including describing and nonreacting to inner experience, and life satisfaction. Serenity also mediated the relation between the mindfulness facet of describing and depressive symptoms. Direct associations were indicated between two mindfulness facets, including observing and nonjudging of inner experience, and depressive symptoms. Taken together, the findings revealed mindfulness facets as major correlates of serenity and psychological outcomes among Chinese meditation practitioners. To foster psychological well-being, researchers, educators, and practitioners should pay attention to role of serenity, describing, and nonreacting to inner experience in mental health.

## Introduction

Mindfulness has been practiced for centuries in the context of Buddhism and contemplative Christianity [1, 2]. Nonetheless, research in this area has only flourished globally over the last two decades [3, 4]. According to Kabat-Zinn [5], mindfulness refers to paying attention nonjudgmentally to present-moment experiences such as emotions, thoughts, and body sensations. Numerous studies have shown that mindfulness has health benefits including better quality of life, better sleep quality, lower stress, fewer anxiety and depressive symptoms, and greater life satisfaction [6, 7, 8, 9, 10, 11]. In addition, mindfulness also fosters greater spirituality. For instance, a meta-analytic study showed that after attending interventions such as mindfulness-based cognitive therapy (MBCT; [12]) and mindfulness-based cancer recovery (MBCR; [13]), participants from diverse clinical and nonclinical samples experienced greater spirituality [14]. As such, studies to date involving diverse international samples have shown that mindfulness is broadly linked to better spirituality, health, and well-being.

### Serenity as a Mediating Mechanism

Spirituality overarches several dimensions, including serenity, which is defined as the spiritual state of inner peace, regardless of the experience of positive and negative life events [15, 16, 17]. More specifically, serenity is a three sub-dimensional construct including acceptance of self and uncontrollable events, ability to tap into the inner haven of peace, and trust in the meaningfulness and profoundness of life [15]. Previous research indicated that higher levels of mindfulness were linked to greater internal peace and harmony [18, 19]. After attending mindfulness-based programs, such as the MBSR [20], health care practitioners reported increased serenity [21, 22, 23]. Importantly, Shapiro et al. [24] theorized that mindfulness practice enables a shift of perspectives that fosters an accepting attitude toward negative emotions, an ability to enter the space of inner calmness, and a recognition of meaningful values, all of which point to greater serenity. Individuals who are more mindful are also more able to experience intense emotions with less reactivity and more objectivity. That is, mindfulness enables individuals to remain calm and non-reactive, despite an awareness of strong emotions [15, 17]. Previous research suggested that the stability of inner peace brought by spirituality was related to positive well-being outcomes, including fewer depressive symptoms and greater life satisfaction [25, 26, 27]. Other studies differentially indicated various aspects of serenity (e.g., acceptance of self and events beyond one’s control, inner peace, and faith in the meaningfulness of life and larger plan) as mediators between mindfulness and well-being outcomes [18, 28, 29]. As a unifying construct, serenity may serve as a mediator between mindfulness facets and well-being outcomes, including depressive symptoms and life satisfaction.

### Differential Contributions of the Facets of Mindfulness

The Five Facets Mindfulness Questionnaires (FFMQ; [30]) is a commonly used measure of mindfulness, encompassing facets including observing, nonjudging of inner experience, describing, acting with awareness, and nonreacting to inner experience. According to Baer et al. [31], observing refers to attending to or noticing experiences internally and externally, namely sounds, emotions, and thoughts. Nonjudging of inner experience refers to adopting a nonevaluative position towards inner experiences, such as feelings and thoughts. Describing involves the labeling of inner experiences using words. Acting with awareness refers to attending to present-moment activities rather than being on autopilot. Finally, nonreacting to inner experience involves the inclination to let inner experiences (e.g., thoughts, feelings) come and go, instead of being taken away by them.

Although the longstanding literature has identified positive associations between mindfulness and well-being outcomes [8, 9, 10], emerging studies have highlighted differential contributions of each mindfulness facet on psychological well-being [32, 33, 34, 35]. For example, a study found that contradictory to other facets, observing was positively associated with psychological distress [32]. The authors speculated that the greater tendency to observe experiences may lead to anxiety or panic, particularly if the experiences are perceived as negative or dangerous. Another study yielding similar findings suggested that observing may be indicative of maladaptive self-attention, which may impede nonjudgmental moment-to-moment awareness and give rise to greater distress [35]. Furthermore, a study involving a nonclinical sample indicated that after controlling for age, gender, and meditation practices, observing and describing did not predict psychological distress, whereas the other three facets of mindfulness, including nonjudging, nonreacting, and acting with awareness, were significant predictors of distress [32]. Baer [36] posited that for individuals without an experience of meditation, observation and attention to the present moment may be impulsive and judgmental. On the contrary, mindfulness practices may enable meditators to observe openly and acceptingly [36]. Therefore, observing may be positively related to well-being outcomes among meditators.

However, a study reported that observing was related to greater rumination and lower attentional control, and that the indirect links between observing and mental/physical health complaints via greater body awareness and nonattachment were significant among samples with or without meditation experience [37]. As such, the association between observing and well-being among meditators and non-meditators remain inconsistent and unclear. As for other mindfulness facets, Bergin and Pakenham [32] reported positive relations between all facets of mindfulness and life satisfaction, except for nonreacting. Given the inconsistency and scarcity of the findings especially among regular meditators, it is crucial to examine the unique contribution of each mindfulness facet in psychological well-being.

### The Present Study

Guided by Shapiro et al.’s [24] theory on mindfulness, the present study investigated serenity as a mediator between mindfulness facets and well-being outcomes, including depressive symptoms and life satisfaction, among mindfulness practitioners. Of note, a majority of research in this area was conducted in general or clinical populations [26, 27, 29, 19].

Relatively few studies have examined the role of mindfulness on mental health among meditators, particularly in diverse Chinese contexts. Among the studies conducted in the Chinese context, findings did show that dispositional mindfulness was linked to depression and life satisfaction [6, 38, 10] and that an aspect of serenity (i.e., self-acceptance) did mediate the association between mindfulness and mental health among emerging adults between 17 and 25 years of age in Hong Kong [25]. Nevertheless, more work is needed to understand the role of the unifying construct of serenity encompassing inner haven of peace, acceptance of self and uncontrollable events, and trust in the meaningfulness and profoundness of life. Based on theory and existing findings, the present study hypothesized that serenity would mediate the association between mindfulness facets and psychological well-being, including depressive symptoms and life satisfaction, among Chinese mindfulness practitioners.

## Materials and Method

### Participants

This study was part of a larger project aiming to investigate the psychological well-being among mindfulness practitioners (Masked for Peer Review). Mindfulness practitioners (*N* = 133; 75.94% female; 24.06% male) were recruited at a three-day transnational meditation event in Hong Kong in 7 to 9 June 2019. Ethics approval was sought by (Granting Body and Protocol Number Masked for Peer Review) prior to the commencement of this study. Written informed consent was collected from participants before they began the self-report questionnaire. Upon completion, the consent forms were collected and stored separately from the questionnaires in locked cabinets, such that participants’ responses were not associated with any person-identifiable information.

All participants were ethnically Chinese who were between 20 and 72 years old (*M* = 47.95 years; *SD* = 11.55 years). Regarding their education, .88% completed primary school, 7.08% completed junior high school, 29.21% completed senior high school, 9.73% had a higher diploma, 28.32% had a bachelor’s degree, 23.01% had a master’s degree, and 1.77% had a doctoral degree. The median monthly household income was HK$30001–$40000 (∼US$3846.28 – $5141.39), which was greater than the median household income in Hong Kong [39].

Regarding religion, 46.62% were Buddhists, 2.26% were non-Buddhist Chinese folk religion believers, 3.76% were Protestants, 2.26% were Catholics, 17.29% were Atheists, 8.27% were agnostic or did not have a particular religious belief, and 19.54% were “other”. Participants reportedly had an average of meditation practice for 2.98 years (*SD* = 4.13 years). In addition, they reported practicing 2.42 hours of meditation each week (*SD* = 2.89 hours).

### Measures

#### Mindfulness

The 20-item Five Facet Mindfulness Questionnaire-Short Form (FFMQ-SF; [40]) was used to assess five facets of mindfulness, including observing, describing, nonjudging, acting with awareness, and nonreacting. Sample items included, “I pay attention to sensations, such as the wind in my hair or sun on my face” (observing), “I’m good at finding words to describe my feelings” (describing), “I tell myself I shouldn’t be feeling the way I’m feeling” (nonjudging), “When I do things, my mind wanders off and I’m easily distracted” (acting with awareness), and “In difficult situations, I can pause without immediately reacting” (nonreacting). Participants rated on a 5-point scale from 1 (*never or very rarely true*) to 5 (*very often or always true*).

Negative items were reverse scored, such that higher average scores indicated greater mindfulness. In this study, Cronbach’s alpha = .83 for observing, .87 for describing, .65 for nonjudging, .89 for acting with awareness, and .88 for nonreacting.

#### Serenity

The 22-item Brief Serenity Scale (BSS; [15]) was used to assess three factors of serenity, including trust, acceptance, and inner haven. Sample items included, “I trust that everything happens as it should” (trust), “In problem situations, I do what I am able to and then accept whatever happens even if I dislike it” (acceptance), and “I experience and inner calm even when I am under pressure” (inner haven). Participants rated on a 5-point scale from 1 (*never*) to 5 (*always*). Item scores were averaged to create three subscale scores. Higher scores indicated greater serenity. In this study, Cronbach’s alphas for the subscales = .91 for acceptance, .94 for inner haven, and .84 for trust.

#### Depressive Symptoms

The 9-item Patient Health Questionnaire (PHQ-9; [41]) was used to assess depressive symptoms over the last 2 weeks on a 4-point scale from 0 (*not at all*) to 3 (*nearly every day*). Sample items included, “Feeling bad about yourself, or that you are a failure or have let yourself or your family down” and “Feeling down, depressed, or hopeless.” Higher averaged scores indicated greater depressive symptoms. In this study, Cronbach’s alpha = .93.

#### Life Satisfaction

The 5-item Satisfaction with Life Scale (SWLS; [42]) was used to assess life satisfaction on a 7-point scale from 1 (*totally disagree*) to 7 (*totally agree*). Sample items included, “I am satisfied with my life” and “So far I have gotten the important things I want in life.” Higher averaged scores indicated greater life satisfaction. In this study, Cronbach’s alpha = .86.

### Data Analysis

A structural equation model (SEM) was conducted using MPLUS, Version 8.3 [43] to investigate the mediating effect of serenity between mindfulness and psychological well-being, including life satisfaction and depressive symptoms, over and above covariates including age, gender, education, household income, experience of meditation, as well as hours of meditation each week. Maximum likelihood method was adopted to investigate the model fit to observed matrices of variance and covariance. Full information maximum likelihood estimation was used to handle missing data at the item or subscale level. Bootstrapping was used to investigate indirect mediation effects.

## Results

Table 1 shows the correlations, means, and *SD*s of the variables. The structural model fit adequately to the data, *χ^2^*(279) = 384.79, *p* < .001, CFI = .94, TLI = .93, RMSEA = .06; SRMR = .06 (see Table 2 and Fig 1 for details). Latent variables including serenity, depressive symptoms, and life satisfaction were significantly indicated by their respective manifest indicators, *p*s < .001. After controlling for age, sex, income, years of education, meditation experience, and hours of meditation per week, two facets of mindfulness including describing and nonreacting were positively associated with serenity (*B* = .16, *SE* = .06, β = .18, *p* = .009 and *B* = .59, *SE* = .06, β = .71, *p* < .001, respectively). Serenity was, in turn, negatively related to depressive symptoms (*B* = -.40, *SE* = .19, β = -.40, *p* = .036) and positively related to life satisfaction (*B* = 1.98, *SE* = .34, β = 1.03, *p* < .001). In addition, two other facets of mindfulness were associated with depressive symptoms. Specifically, observing was positively related to depressive symptoms (*B* = .21, *SE* = .07, β = .28, *p* = .004), whereas nonjudging was negatively related to depressive symptoms (*B* = -.23, *SE* = .07, β = -.26, *p* = .001).

**Fig 1.**
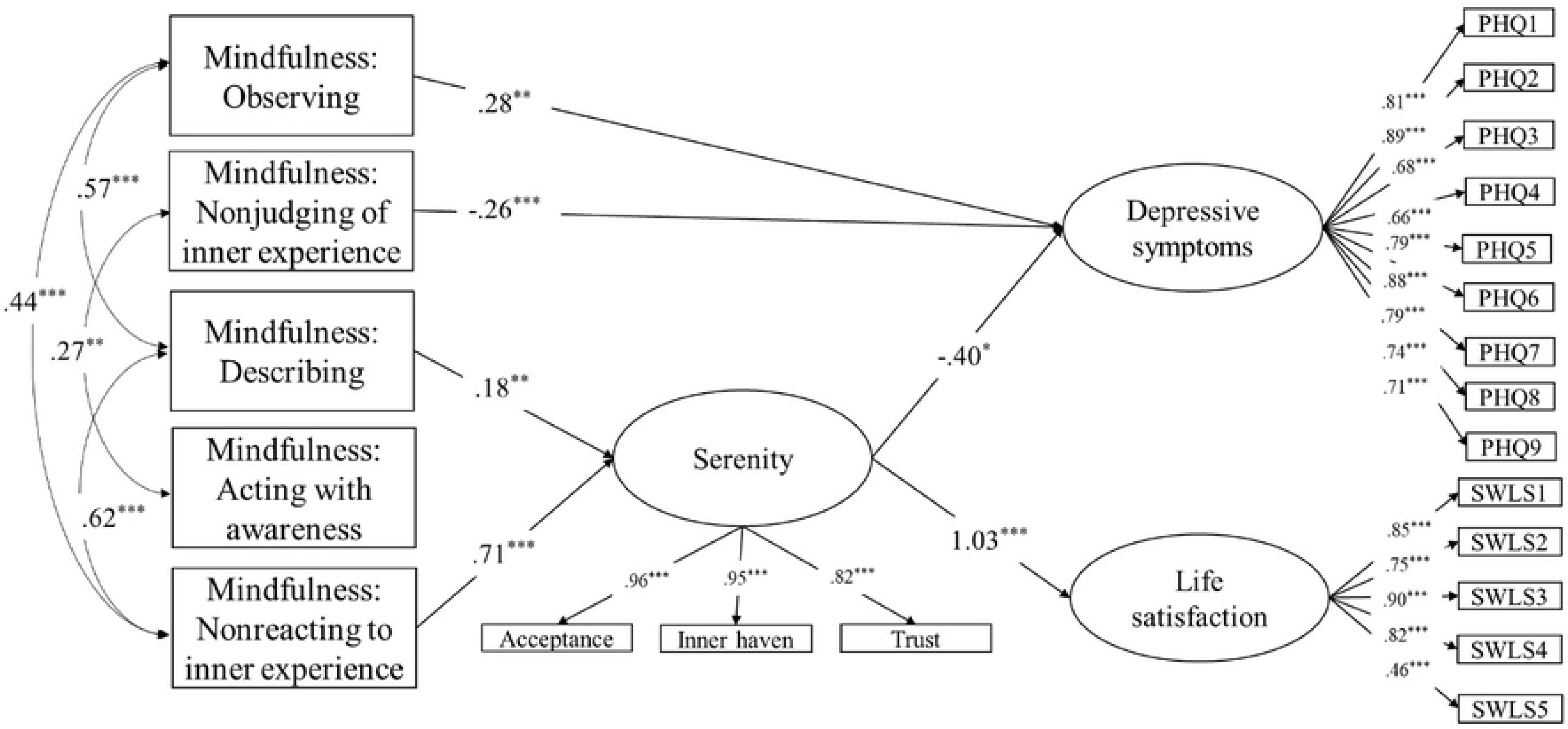
Final model of serenity as a mediator between mindfulness and depressive symptoms and life satisfaction.

**Table 1.** Zero-order Correlations, Means, and Standard Deviations of the Variable.

| Variable | 1 | 2 | 3 | 4 | 5 | 6 | 7 | 8 | 9 | 10 | 11 | 12 | 13 | 14 | 15 | 16 |
| --- | --- | --- | --- | --- | --- | --- | --- | --- | --- | --- | --- | --- | --- | --- | --- | --- |
| 1. Sex (0 = male; 1 = female) | — |  |  |  |  |  |  |  |  |  |  |  |  |  |  |  |
| 2. Age | -.05 | — |  |  |  |  |  |  |  |  |  |  |  |  |  |  |
| 3. Education | -.00 | -.37*** | — |  |  |  |  |  |  |  |  |  |  |  |  |  |
| 4. Income | .06 | -.02 | .20* | — |  |  |  |  |  |  |  |  |  |  |  |  |
| 5. Experience of meditation (in months) | -.02 | .30** | .06 | -.09 | — |  |  |  |  |  |  |  |  |  |  |  |
| 6. Hours of meditation per week | -.02 | .13 | -.09 | -.09 | .40*** | — |  |  |  |  |  |  |  |  |  |  |
| 7. Mindfulness: Observing | -.04 | .05 | .05 | -.10 | .15 | .23* | — |  |  |  |  |  |  |  |  |  |
| 8. Mindfulness: Describing | -.11 | .20* | .17 | .17 | .21* | .09 | .58*** | — |  |  |  |  |  |  |  |  |
| 9. Mindfulness: Acting with awareness | -.12 | -.02 | -.05 | .14 | .07 | .10 | -.01 | -.10 | — |  |  |  |  |  |  |  |
| 10. Mindfulness: Nonjudging | -.03 | .11 | -.13 | -.01 | -.07 | .04 | -.15 | -.12 | .27** | — |  |  |  |  |  |  |
| 11. Mindfulness: Nonreacting | -.18 | .25** | -.02 | .05 | .34*** | .34*** | .45*** | .62*** | .02 | .01 | — |  |  |  |  |  |
| 12. Serenity: Acceptance | -.17 | .21* | .09 | .12 | .32*** | .31** | .42*** | .63*** | .02 | -.01 | .83*** | — |  |  |  |  |
| 13. Serenity: Inner haven | -.13 | .22* | .05 | .08 | .36*** | .35*** | .48*** | .65*** | -.02 | -.03 | .84*** | .90*** | — |  |  |  |
| 14. Serenity: Trust | -.20* | .19* | .15 | .14 | .27** | .23* | .40*** | .57*** | -.00 | -.05 | .70*** | .82*** | .75*** | — |  |  |
| 15. Depressive symptoms | .08 | -.35*** | .03 | -.15 | -.18 | -.19 | .05 | -.21* | -.18* | -.31*** | -.39*** | -.39*** | -.37*** | -.30** | — |  |
| 16. Life satisfaction | .02 | .23* | .12 | .22* | .21* | .08 | .30** | .51*** | .00 | -.06 | .62*** | .71*** | .72*** | .62*** | -.36*** | — |
| *M* | — | 49.95 | 5.72 | 4.59 | 35.79 | 2.42 | 3.44 | 3.22 | 3.22 | 3.00 | 3.21 | 3.46 | 3.38 | 3.49 | .60 | 4.78 |
| *SD* | — | 11.55 | 2.17 | 2.86 | 49.60 | 2.59 | .91 | .81 | 1.05 | .78 | .84 | .72 | .83 | .82 | .66 | 1.30 |

Note. \* $p < .05$ , \*\* $p < .01$ , \*\*\* $p < .001$ .

**Table 2.** Path Estimates of the Mediation Model.

| Parameter | Unstandardized B (SE) | Standardized $\beta$ |
| --- | --- | --- |
| **Measurement Model** |  |  |
| Serenity |  |  |
| → Acceptance | 1.00<sup>f</sup> | .96*** |
| → Inner haven | 1.15 (.07) | .95*** |
| → Trust | .98 (.07) | .82*** |
| Depressive symptoms |  |  |
| → PHQ item 1 | 1.00<sup>f</sup> | .81*** |
| → PHQ item 2 | 1.04 (.09) | .89*** |
| → PHQ item 3 | .91 (.11) | .68*** |
| → PHQ item 4 | .84 (.11) | .66*** |
| → PHQ item 5 | .92 (.09) | .79*** |
| → PHQ item 6 | 1.22 (.11) | .88*** |
| → PHQ item 7 | .98 (.10) | .79*** |
| → PHQ item 8 | .88 (.10) | .74*** |
| → PHQ item 9 | .60 (.07) | .71*** |
| Life satisfaction |  |  |
| → SWLS item 1 | 1.00<sup>f</sup> | .85*** |
| → SWLS item 2 | .82 (.09) | .75*** |
| → SWLS item 3 | 1.07 (.09) | .90*** |
| → SWLS item 4 | 1.04 (.10) | .82*** |
| → SWLS item 5 | .63 (.13) | .46*** |
| **Structural Model** |  |  |
| Mindfulness: Observing |  |  |
| → Serenity | .03 (.05) | .04 |
| → Depressive symptoms | .21 (.07) | .28** |
| → Life satisfaction | -.06 (.11) | -.04 |
| Mindfulness: Describing |  |  |
| → Serenity | .16 (.06) | .18** |
| → Depressive symptoms | -.02 (.10) | -.03 |
| → Life satisfaction | -.08 (.16) | -.05 |
| Mindfulness: Acting with awareness |  |  |
| → Serenity | .01 (.03) | .01 |
| → Depressive symptoms | -.09 (.05) | -.13 |
| → Life satisfaction | .07 (.08) | .06 |
| Mindfulness: Nonjudging |  |  |
| → Serenity | -.01 (.04) | -.01 |
| → Depressive symptoms | -.23 (.07) | -.26** |
| → Life satisfaction | -.08 (.11) | -.05 |
| Mindfulness: Nonreacting |  |  |
| → Serenity | .59 (.06) | .71*** |
| → Depressive symptoms | -.06 (.14) | -.07 |
| → Life satisfaction | -.19 (.23) | -.12 |
| Serenity |  |  |
| → Depressive symptoms | -.40 (.19) | -.40* |
| → Life satisfaction | 1.98 (.34) | 1.03*** |
| **Covariance and Error Covariance** |  |  |
| Mindfulness: Observing |  |  |
| ↔ Mindfulness: Describing | .41 (.08) | .57*** |
| ↔ Mindfulness: Acting with awareness | -.01 (.09) | -.01 |
| ↔ Mindfulness: Nonjudging | -.11 (.07) | -.15 |
| ↔ Mindfulness: Nonreacting | .33 (.07) | .44*** |
| Mindfulness: Describing |  |  |
| ↔ Mindfulness: Acting with awareness | -.08 (.08) | -.10 |
| ↔ Mindfulness: Nonjudging | -.07 (.06) | -.11 |
| ↔ Mindfulness: Nonreacting | .41 (.07) | .62*** |
| Mindfulness: Acting with awareness |  |  |
| ↔ Mindfulness: Nonjudging | .22 (.08) | .27** |
| ↔ Mindfulness: Nonreacting | .02 (.08) | .02 |
| Mindfulness: Nonjudging |  |  |
| ↔ Mindfulness: Nonreacting | .01 (.06) | .01 |
| Sex (0 = male; 1 = female) |  |  |
| ↔ Mindfulness: Observing | -.02 (.03) | -.08 |
| ↔ Mindfulness: Describing | -.04 (.02) | -.15 |
| ↔ Mindfulness: Acting with awareness | -.02 (.03) | -.07 |
| ↔ Mindfulness: Nonjudging | .00 (.02) | .01 |
| ↔ Mindfulness: Nonreacting | -.06 (.02) | -.21* |
| → Serenity | -.01 (.11) | -.01 |
| → Depressive symptoms | -.06 (.17) | -.03 |
| → Life satisfaction | .54 (.27) | .13* |
| Age |  |  |
| ↔ Mindfulness: Observing | .30 (1.03) | .03 |
| ↔ Mindfulness: Describing | 1.85 (.93) | .20* |
| ↔ Mindfulness: Acting with awareness | -.53 (1.20) | -.04 |
| ↔ Mindfulness: Nonjudging | .73 (.92) | .08 |
| ↔ Mindfulness: Nonreacting | 2.03 (.93) | .21* |
| → Serenity | .00 (.00) | .02 |
| → Depressive symptoms | -.02 (.01) | -.26** |
| → Life satisfaction | .01 (.01) | .12 |
| Education level |  |  |
| ↔ Mindfulness: Observing | .19 (.27) | .07 |
| ↔ Mindfulness: Describing | .42 (.24) | .17 |
| ↔ Mindfulness: Acting with awareness | -.14 (.31) | -.04 |
| ↔ Mindfulness: Nonjudging | -.31 (.24) | -.13 |
| ↔ Mindfulness: Nonreacting | .02 (.23) | .01 |
| → Serenity | .02 (.01) | .07 |
| → Depressive symptoms | -.01 (.02) | -.05 |
| → Life satisfaction | .01 (.03) | .03 |
| Income |  |  |
| ↔ Mindfulness: Observing | -.22 (.27) | -.08 |
| ↔ Mindfulness: Describing | .41 (.22) | .18* |
| ↔ Mindfulness: Acting with awareness | .45 (.30) | .15 |
| ↔ Mindfulness: Nonjudging | .02 (.22) | .01 |
| ↔ Mindfulness: Nonreacting | .17 (.23) | .07 |
| → Serenity | -.01 (.01) | .04 |
| → Depressive symptoms | -.01 (.02) | -.06 |
| → Life satisfaction | .03 (.03) | .06 |
| Experience of meditation (in months) |  |  |
| ↔ Mindfulness: Observing | 7.20 (4.35) | .16 |
| ↔ Mindfulness: Describing | 8.13 (3.77) | .21* |
| ↔ Mindfulness: Acting with awareness | 1.86 (5.08) | .04 |
| ↔ Mindfulness: Nonjudging | -3.70 (3.78) | -.10 |
| ↔ Mindfulness: Nonreacting | 13.29 (4.02) | .33*** |
| → Serenity | .00 (.00) | .04 |
| → Depressive symptoms | .00 (.00) | .00 |
| → Life satisfaction | -.00 (.00) | .02 |
| Hours of meditation per week |  |  |
| ↔ Mindfulness: Observing | .64 (.27) | .25* |
| ↔ Mindfulness: Describing | .23 (.22) | .10 |
| ↔ Mindfulness: Acting with awareness | .25 (.30) | .08 |
| ↔ Mindfulness: Nonjudging | -.01 (.22) | -.00 |
| ↔ Mindfulness: Nonreacting | .76 (.23) | .32*** |
| → Serenity | .01 (.02) | .06 |
| → Depressive symptoms | -.01 (.01) | -.06 |
| → Life satisfaction | -.08 (.02) | -.18* |
| Depressive symptoms ↔ Life satisfaction | -.08 (.04) | -.23 |

Note. \*p < .05, \*\*p < .01, \*\*\*p < .001.

Based on the above findings, the indirect effects between two facets of mindfulness, including describing and nonreacting, and depressive symptoms and life satisfaction through serenity were investigated. Based on 5000 bootstrap samples with replacement, the 95% confidence interval (CI) indicated that the unstandardized indirect effect between describing and depressive symptoms did not include a zero (CI: [-.21, -.001]). However, the 95% CI indicated that the unstandardized indirect effect between nonreacting and depressive symptoms included a zero (CI: [-.56, .04]). As for life satisfaction, based on 5000 bootstrap samples with replacement, the 95% confidence interval (CI) indicated that the unstandardized indirect effect between describing and life satisfaction did not include a zero (CI: [.06, .36]). In addition, the 95% CI indicated that the unstandardized indirect effect between nonreacting and life satisfaction did not include a zero (CI: [.46, 1.11]). Hence, serenity mediated between describing and life satisfaction. Serenity also mediated between two facets of mindfulness, including describing and nonreacting, and life satisfaction.

## Discussion

Guided by Shapiro et al.’s [24] theory on mindfulness, this study examined serenity as a mediating process between mindfulness facets and psychological outcomes. The present findings provided incremental support to suggest serenity as a mediator between two facets of mindfulness [30], namely describing and nonreacting to inner experience, and life satisfaction.

Serenity also mediated the relation between describing and depressive symptoms. In addition, direct associations were evidenced between mindfulness facets (i.e., observing and nonjudging of inner experience) and depressive symptoms. Altogether, the findings revealed mindfulness facets as major correlates of serenity and psychological outcomes among Chinese meditation practitioners.

In this study, serenity was associated with mindfulness facets (i.e., describing and nonreacting to inner experience) and psychological well-being. When people were capable of describing clearly and responding skillfully (vs. reacting mindlessly) to their experiences, they were also more likely to have greater serenity, e.g., accept themselves and uncontrollable events, touch the inner haven of peace, and trust that life is meaningful and profound [15]. Between the two mindfulness facets, the strength of association between nonreacting to inner experience and serenity was particularly strong, suggesting that the ability to pause, to “step back” without being taken over by inner experience, and to let go were especially crucial to shifting perspectives that foster serenity (see also [24]). Hence, to promote serenity, researchers, educators, and mindfulness practitioners should pay close attention to role of nonreacting to inner experience.

Somewhat surprisingly, other facets of mindfulness including observing, nonjudging of inner experience, and acting with awareness were unrelated to the spiritual state of serenity. Even though the mindfulness facets were intercorrelated, their relations with serenity did not bear out after describing and nonreacting to inner experience were taken into account. Nonjudging of inner experience and observing were, instead, directly related to depressive symptoms, a core indicator of mental health. Consistent with previous research [44, 33, 45], when people were more judgmental and critical of their inner experience (e.g., “my thoughts are good, bad, or inappropriate”) and of how they should be (e.g., “I shouldn’t feel this way”), they were more likely to have greater depressive symptoms. Nevertheless, when people were more observing of their experiences internally and externally, they were also more likely to have greater depressive symptoms. The findings corroborated previous research [32, 35] to suggest that observing was positively related to depressive symptoms. As speculated by Tran et al. [35], perhaps observing was indicative of maladaptive self-attention that gave rise to depressive symptoms. Although Baer [36] argued that people without meditation experiences might observe the present judgmentally and impulsively, whereas practitioners might observe with openness and acceptance, in this study we found a positive link between observing and depressive symptoms among practitioners with a regular meditation practice (See [46] and [37] for similar findings). In addition, our SEM findings revealed that observing was not related to nonjudging of inner experience at all (see Fig 1). Given that observing was moderately related to describing (*r* = .57, *p* < .001) and nonreacting to inner experience (*r* = .44, *p* < .001), the finding between observing and depressive symptoms could also be the result of multicollinearity. Hence, the study of observing and depression among people with and without meditation experience warrants future investigation.

Interestingly, acting with awareness was not related to serenity, depressive symptoms, or life satisfaction. That is, the ability to pay attention without being easily distracted, daydreaming, or worrying was not related to serenity and psychological functioning, particularly when other mindfulness facets were taken into account. The null finding was intriguing, as it diverged from existing findings between mindful attention awareness [47], mind wandering, and psychological well-being [48, 49]. It also contradicted previous findings between mindful awareness and serenity based on samples from the Western contexts [15, 27]. To fully understand the link between acting with awareness and serenity, or the lack thereof, researchers should pay a closer attention to the use of assessments (e.g., FFMQ; [30] vs. Mindfulness Attention and Awareness Scale, MAAS; [47]), the role of culture (e.g., collectivistic vs. individualistic cultures), and sample characteristics (e.g., people with and without experiences of meditation, clinical samples, non-clinical samples).

Consistent with previous findings [15, 27], the core aspects of serenity were related to both depressive symptoms and life satisfaction. These findings suggested an important link between spiritual health and psychological well-being. In addition, serenity served as a potential mechanism between the mindfulness facets of describing and nonreacting to inner experience and psychological well-being. Supporting the literature, the findings inform researchers, educators, and mindfulness and mental health practitioners of the relevance of being mindful and serene in mental health.

## Limitations and Future Directions

The present study has several limitations. First of all, the utilization of a self-report questionnaire could have resulted in method bias [50]. Thus, future studies should include multiple methods and multiple informants to reduce potential biases. Second, due to the cross-sectional design of the present study, we were unable to draw conclusions on the temporal sequence between the study variables. Future research should utilize a longitudinal design in testing mediation [51]. Third, although nonreacting to inner experience and aspects of serenity are theoretically distinct variables, their zero-order correlations were large [52], ranging between *r*s = .70 and .84 (see Table 1). According to Baer et al. [31 p. 330], nonreacting to inner experience refers to “the tendency to allow thoughts and feelings to come and go, without getting caught up in or carried away by them.” As an overarching construct, serenity refers to the spiritual state of inner peace, regardless of the experience of positive and negative life events [15]. Although both constructs suggest nonreactivity to experiences, the mindfulness facet of nonreacting points primarily to inner experiences (e.g., thoughts, feelings), whereas serenity involves spiritual states regardless of internal vs. external experiences or stimuli. Serenity further involves an added layer of peace, trust, and acceptance beyond nonreactivity. Despite the potential differences, future research should distinguish or clarify theoretically and empirically the relation between non-reactivity and serenity. Finally, in this study, the recruitment of mindfulness practitioners took place in a meditation event and most of the study participants were women. Hence, future work is necessary to increase generalizability.

Despite the above limitations, the present study lends preliminary support to Shapiro et al.’s [24] theory on mindfulness and broadens the literature involving serenity as a process between mindfulness facets and psychological outcomes. As practical implications, educators and mental health practitioners should attend to the relations between various facets of mindfulness, serenity, and mental health. In terms of research implications, theoretical and empirical studies are necessary to clarify why, how, and when mindfulness facets are linked to serenity and mental health outcomes. Translational research gearing towards enhancing mindfulness and serenity also merits future investigations.

## Data Availability

All relevant data are within the manuscript and its Supporting Information files. The files do not contain participants' identifying information.

